# Computations of State Ventilation and Respiratory Parameters

**DOI:** 10.1101/2021.11.08.21266078

**Authors:** Quangang Yang

## Abstract

**Background:** In mechanical ventilation, there are still some challenges to turn a modern ventilator into a fully reactive device, such as lack of a comprehensive target variable and the unbridged gap between input parameters and output results. This paper aims to present a state ventilation which can provide a measure of two primary, but heterogenous, ventilation support goals. The paper also tries to develop a method to compute, rather than estimate, respiratory parameters to obtain the underlying causal information.

**Methods:** This paper presents a state ventilation, which is calculated based on minute ventilation and blood gas partial pressures, to evaluate the efficacy of ventilation support and indicate disease progression. Through mathematical analysis, formulae are derived to compute dead space volume/ventilation, alveolar ventilation, and CO2 production.

**Results:** Measurements from a reported clinical study are used to verify the analysis and demonstrate the application of derived formulae. The state ventilation gives the expected trend to show patient status, and the calculated mean values of dead space volume, alveolar ventilation, and CO2 production are 158mL, 8.8L/m, and 0.45L/m respectively for a group of patients.

**Discussions and Conclusions:** State ventilation can be used as a target variable since it reflects patient respiratory effort and gas exchange. The derived formulas provide a means to accurately and continuously compute respiratory parameters using routinely available measurements to characterize the impact of different contributing factors.

## INTRODUCTION

Ventilation support is a clinical intervention used to sustain respiration in patients who are unable to breathe on their own or find it difficult to do so. Mechanical ventilation, the most common ventilatory support, is widely used for treating respiratory failure or deficiency. During the past decades, improvements in our understanding of respiratory physiology and pathology, as well as various technological advancements, have allowed the development of ventilators that are more sophisticated and versatile [1]. These have more ventilation modes, expanded applications, and can provide patients with more accurate and desired ventilation.

The ventilator is still evolving, and some researchers have given insight into what future ventilators will look like [2,3]. The application of Artificial Intelligence (AI) has gained a huge amount of interest in healthcare due to technological advancements in machine learning and data science. Because of the large amount of information that mechanical ventilation can collect and the expertise it requires in decision making, it should be a strong candidate for AI application. Some recent AI applications in mechanical ventilation include data acquisition [4], weaning management [5], predicting the need for ventilation support [6], and optimizing ventilation settings [7]. However, there are still major barriers preventing a ventilator from becoming a fully reactive machine even though most modern ventilators have networking, data mining, and computation capabilities. There is a lack of a comprehensive target variable which can reflect the two primary goals of ventilation support: unloading respiratory effort and improving pulmonary gas exchange. Although current AI can recommend ventilatory settings for a patient based on historical data [7], without a comprehensive target variable, it is nearly impossible for a ventilator to tune the settings on the fly in response to changes in the patient’s condition. Recently, a ventilatory ratio was proposed to measure ventilatory efficiency [8,9]. It took minute ventilation and partial CO_2_ pressure into consideration, but blood oxygenation was not included. Another challenge is that the information collected by present ventilators is generally fragmented and of little use to AI application technologies alike. The underlying causal information, such as CO_2_ production, is not readily available to characterize the impact of different factors. There is still a gap between input parameters, such as pressure, and output results, such as a patient’s blood gas level. For instance, it is hard to tell if a better blood gas outcome under pressure support is a result of unloaded respiratory effort or increased alveolar ventilation. Recent studies on lung characteristics have mostly focused on lung mechanics such as compliance [10], but have rarely included other aspects, such as dead space and carbon dioxide production. Patients’ dead space is often estimated in clinical practice through considerations of weight or height, as other methods, such as Bohr’s method, are impractical to implement during therapy. For example, dead space volume is estimated by inputting a patient’s height into ResMed’s iVAPS ventilation mode. However, the estimation concerns anatomic dead space, rather than physiologic dead space, and the actual dead space may vary during treatment. Consequently, a patient’s alveolar ventilation cannot be accurately and continuously computed.

## METHODS

The state ventilation *SV* under mechanical ventilation can be defined as the product of minute ventilation 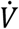 and the ratio of partial CO_2_ pressure 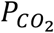 to partial O_2_ pressure 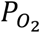,

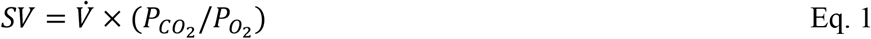

Both 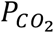 and 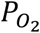 are routine measurements, but to enable real-time calculation, a ventilator must either be able to measure them directly or connect to external devices for measurements. A pulse oximetry provides an easy and reasonably accurate way to measure blood oxygenation while a transcutaneous SenTec™ device can be used to get both *PtcCO*_*2*_, and *SpO*_2_. *PtcCO*_*2*_ can provide an accurate surrogate for 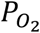 [11,12], and *SpO*_2_ can be used to derive 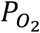. *SV* has a unit of litre per minute and is thus called ventilation here.

Introducing a state index, *SI*, as the product of minute ventilation and CO_2_ partial pressure to have:

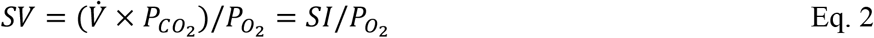

where

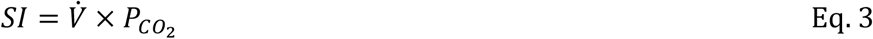

Minute ventilation is the sum of alveolar ventilation and dead space ventilation:

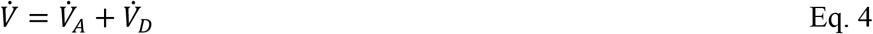

where 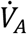 is alveolar ventilation and 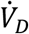 is dead space ventilation. Thus, *SI* can be written as:

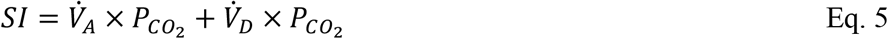

Alveolar ventilation equation is expressed as [13]:

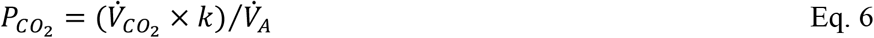

where *k* is the conversion constant and 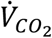 is the CO_2_ removed by the lung, which equals the CO_2_ produced from metabolism in the steady state condition. Substituting Eq. 6 into Eq. 5, and rearranging the equation, we get:

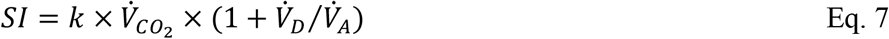

This equation suggests that the state index *SI* is directly proportional to the patient’s CO_2_ production, which reflects the respiratory effort, and related to the dead space/alveolar ventilation ratio, which has a huge impact on gas exchange.

Substituting Eq. 3 into Eq. 7:

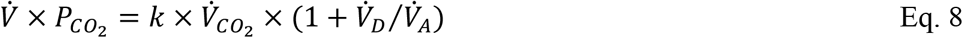

To simplify further analysis, assuming a patient is in a stable condition and there is no big variation in respiratory rate, both dead space ventilation 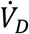 and CO_2_ production 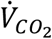 can thus be treated as constant. A minor change in pressure support or tidal volume will result in a small minute ventilation change and consequently, blood CO_2_ level, but will have negligible effect on dead space and CO_2_ production. Differentiate both sides of Eq. 8:

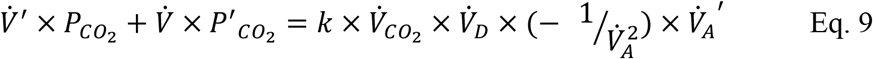

To avoid confusion, it is worth to point out that the dot carried out by a term has a meaning of “minute”, such as minute ventilation 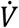, while a prime indicates a derivative with time. Again, with constant 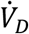, differentiate Eq. 4 to get:

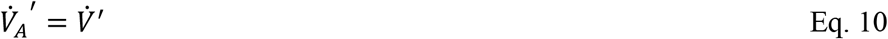

And Eq. 6 can be rewritten as:

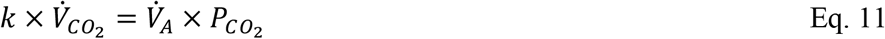

Substituting Eq. 10 and Eq. 11 into the right-hand side of Eq. 9, and rearranging the equation to have:

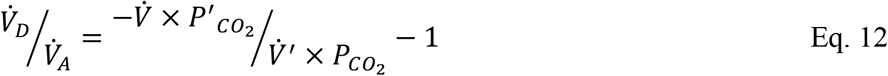

If both minute ventilation 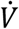 and partial CO_2_ pressure 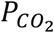 are continuously measured, their gradients, 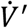 and 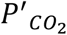, can be easily derived. Therefore, the ratio of dead space ventilation over alveolar ventilation can be computed from Eq. 12.

At the first glance, the right-hand side of Eq. 12 will give a negative value. However, minute ventilation and partial CO_2_ pressure normally change in the opposite directions, suggesting that 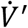 and 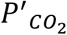 will have opposite signs. Hence the first term on the right-hand side is positive and should be greater than 1.

Solving Eq. 4 and Eq. 12 simultaneously to get alveolar ventilation and dead space ventilation:

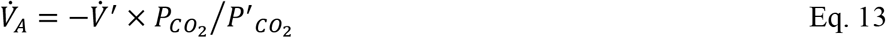

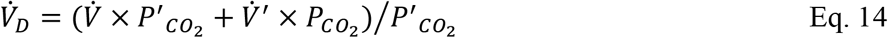

Substituting Eq. 12 into Eq. 8 to obtain CO_2_ production:

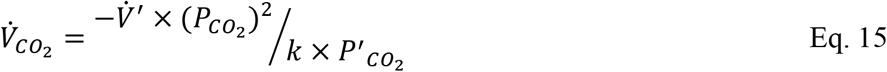

With these equations, alveolar and dead space ventilation, and CO2 production can be computed continuously or at any selected point. Dead space volume can be easily obtained when 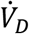 is known:

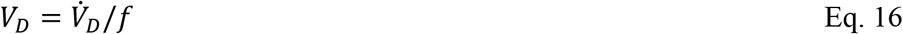

where *f* is the respiratory rate.

## RESULTS

Reported data in existing literature will be used to demonstrate a numerical analysis since the author does not have the resources to perform experimental verification. In 2013, Briones Claudett *et al* reported a study to compare the use of two ventilatory support strategies in patients with Chronic Obstructive Pulmonary Disease (COPD) and hypercapnic encephalopathy upon immediate arrival at the emergency department/ICU [14]. A total of 22 patients were recruited and evenly divided into two groups: the experimental group received Bilevel Positive Airway Pressure-Spontaneous/Timed (BiPAP S/T) with Average Volume Assured Pressure Support (AVAPS), and the control group received conventional BiPAP S/T. In their study, patients’ arterial blood gases were measured at the beginning, and after 1 hour, 3 hours, and 12 hours of ventilatory support. Some of the measurements, as listed in Table 1, will be used here to verify the concept of state ventilation and demonstrate the computations of respiratory parameters.

**Table 1:**
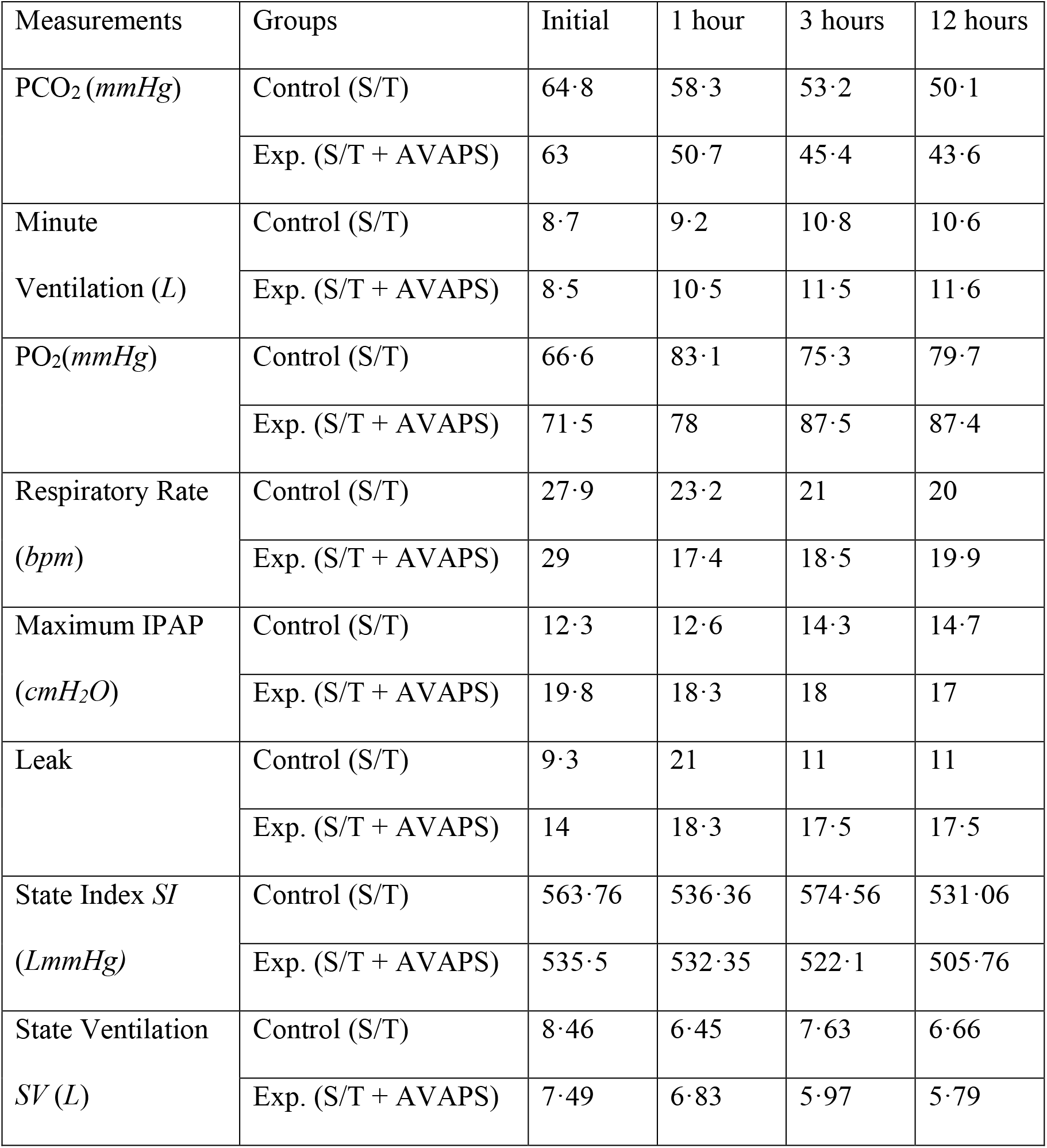
Some measurements from Claudett’s study and state ventilations and indexes.

### State Ventilation and Index

Figure 1 graphically shows the state ventilation trends of the two groups, and their state indexes are shown in Figure 2 for comparison. Both the state ventilation and index of the experimental group show the expected behaviour, decreasing continuously over time, while they experience a large drop at 1 hour for the control group and followed by a jump at 3 hours.

**Figure 1:**
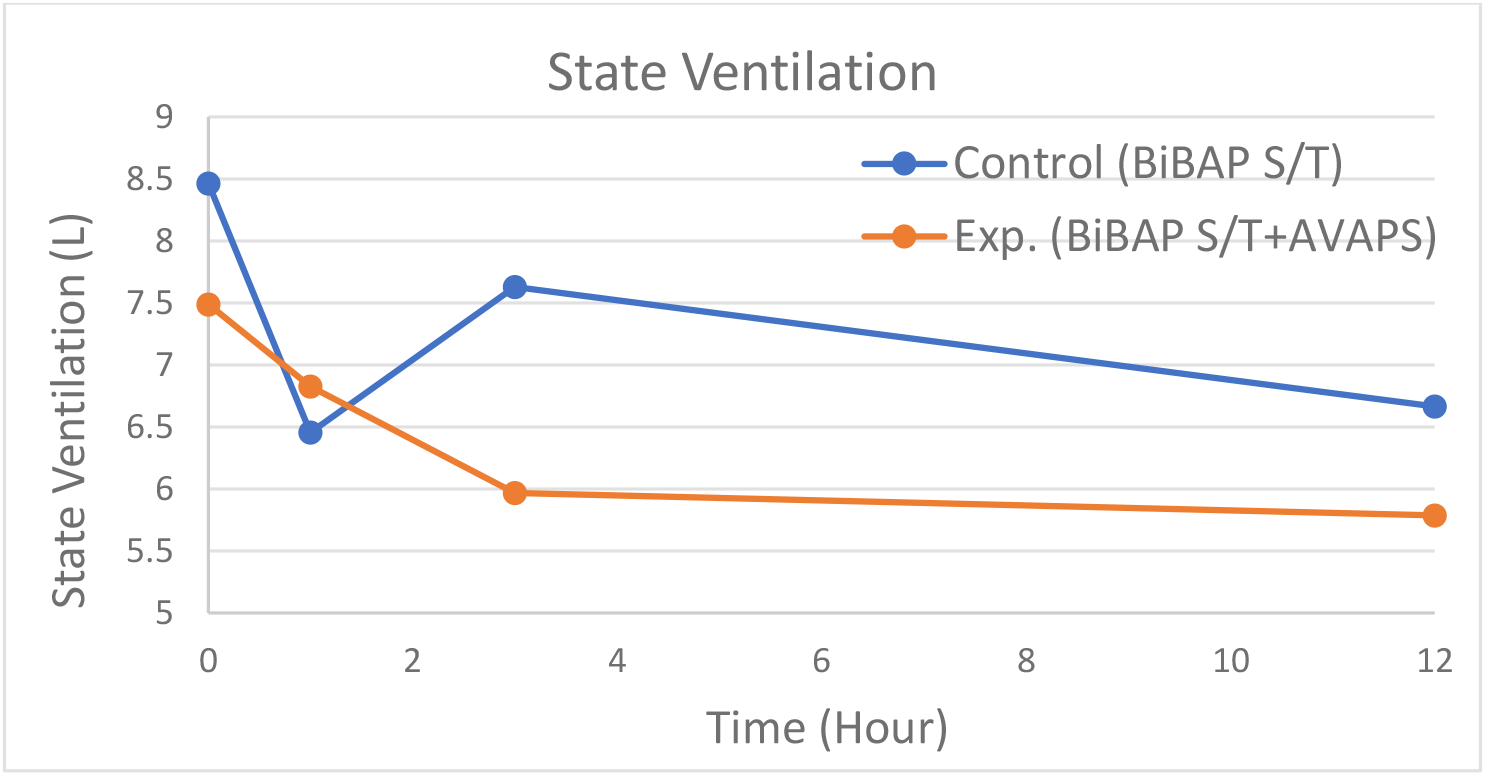
State ventilation over time of the two groups.

**Figure 2:**
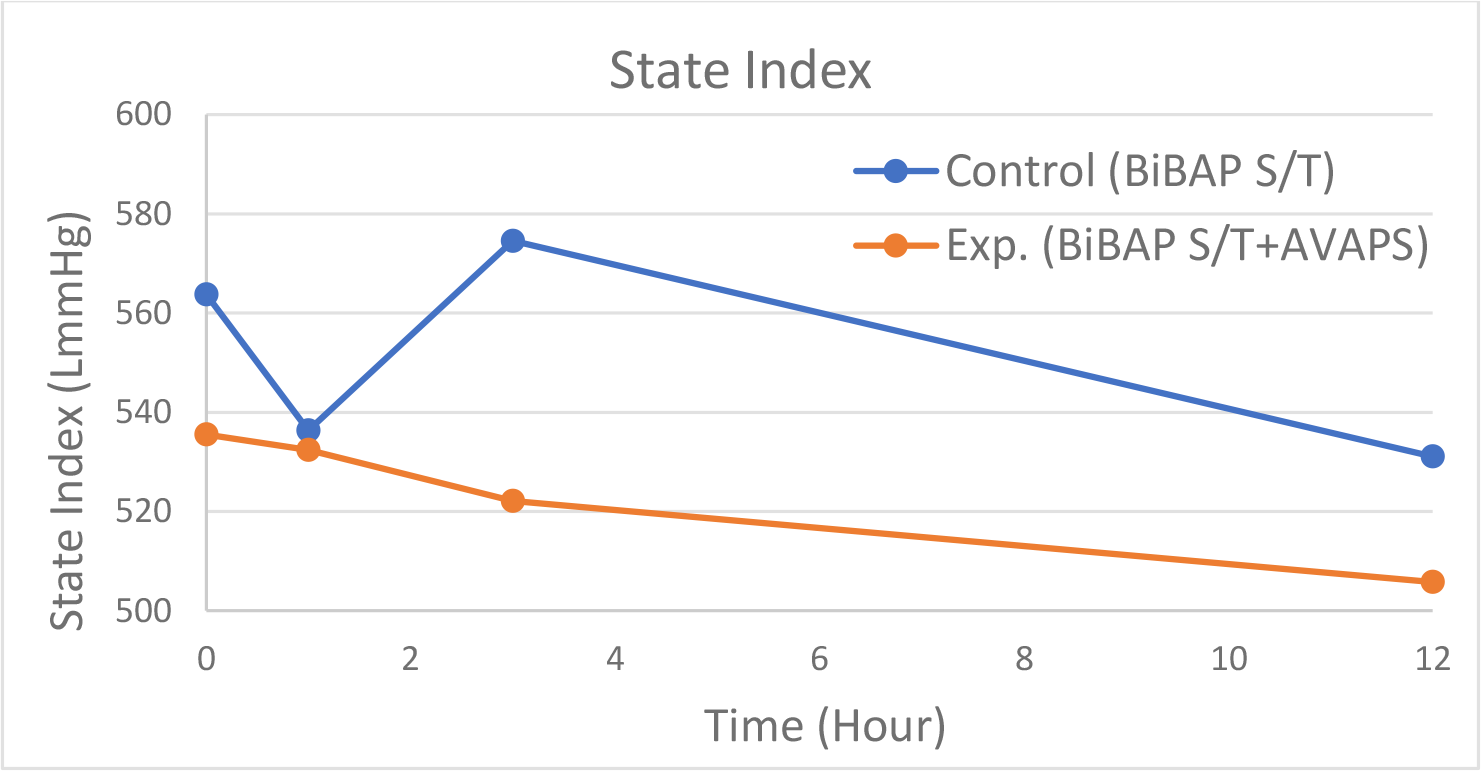
State index over time of the two groups.

Though it is extremely hard to understand the exact cause of this fluctuation due to a lack of underlying information, such as alveolar ventilation and CO_2_ production, we may still be able to make a few comments by crosschecking the data in Table 1:

- Initially, the state ventilation and index of the control group are 0·97*L* and 28·26LmmHg higher than that of the experimental group, indicating that this group has higher CO_2_ production 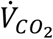 and/or a larger 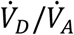 ratio as suggested in Eq. 7. Considering that the control group has a 1·1bpm lower respiratory rate and a 0·2*L* higher minute ventilation compared to the experimental group, the larger 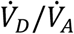 if present, is caused by a greater dead space volume *V*_*D*_.
- Compared with a state ventilation drop of 0·66*L* for the experimental group, the control group has a far larger drop of 2·01*L* in the first hour, suggesting that the conventional BiPAP S/T used in this group is initially more effective. The control group only experiences a minute ventilation increase of 0·5L and a respiratory rate decrease of 4·7bpm, whereas these two values are 2L and 11·6bpm respectively for the experimental group. Thus, it is believed that the control group has a greater reduction of dead space ventilation due not only to the decreased respiratory rate but also to a large reduction in dead space volume. As a result, the control group has a 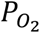increase of 16·5mmHg, 10mmHg more than the experimental group does.
- At 3 hours, the experimental group continues to improve, but the control group has a state ventilation jump. Generally, the fluctuation for an individual patient may suggest an unstable condition, or a circuit leakage resulting in inaccurate minute ventilation. However, the leak shown in Table 1 is minimal, and given the group size of 11 patients, it is almost impossible that every patient is unstable over such a long period. The minute ventilation of the control group increases by 1·6*L* from Hour 1 to Hour 3 because the IPAP in this group is 1·7*cmH2O* higher. However, this increase is not effective in terms of gas exchange as 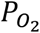 decreases by 7·8*mmHg*, though it results in a lower 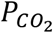. A possible explanation for this phenomenon is that the increased lung volume causes a higher vascular resistance due to the reduced capillary calibre. Consequently, the ventilation-perfusion inequality is worse and the 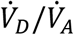 ratio is thus higher, ultimately resulting in a higher state ventilation.
- At 12 hours, both groups keep improving and become stable. The drop in the state ventilation from the beginning to this time is 1·8*L* for the control group, and 1·7*L* for the experimental group, suggesting that BiPAP S/T performs slightly better over the 12 hour period.

### Computation of Respiratory Parameters

After examining Table 1, we use the data of the experimental group at 3 hours to demonstrate the computations of 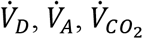, and *V*_*D*_ as the patients in this group have relatively stable conditions, indicated by the mean respiratory rate increase of only 1·1 bpm within a relatively short time of 2 hours. To obtain the derivatives 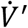 and 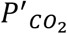 the change of both minute ventilation and partial CO_2_ pressure are assumed to be linear from 1 hour to 3 hours, so that 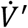 and 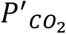 can be computed as:

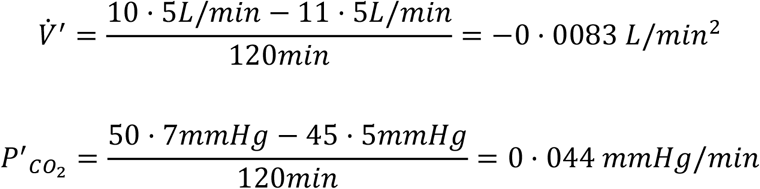

Then, utilizing Eq. 13-14:

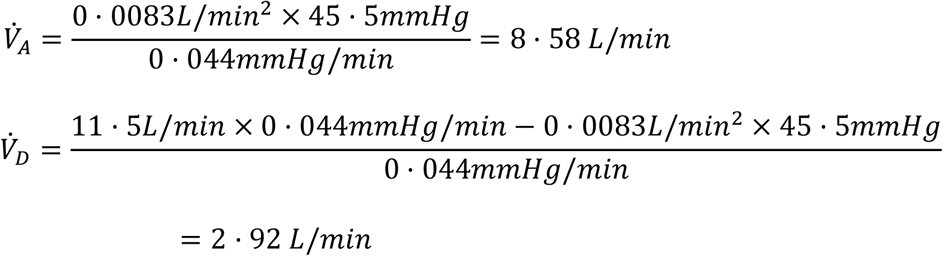

Using a *K* value of 863mmHg in Eq. 15 to get:

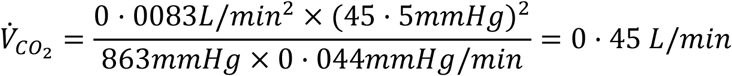

The dead space volume is calculated as:

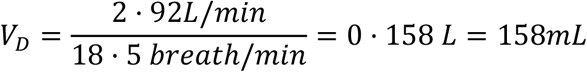

These values are in the ballpark. It should be pointed out again that these are the mean values of the experimental group patients, and the computation accuracy is impacted by the linear assumption of 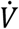 and 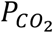 over time from 1 hour to 3 hours.

## DISCUSSIONS

As shown in Figure 1 and 2, the state ventilation and index behave similarly, but the state ventilation curves are generally smoother for both groups. The difference between them comes from the fact that alveolar dead space, or alveolar ventilation, plays a larger role on 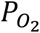 than on 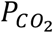 [15]. The state ventilation, thus, presents a clearer and more complete picture as it also reflects blood oxygenation. For example, in a hypoventilation patient with a high 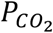 due to reduced ventilation, the state index may only show a small difference whilst the state ventilation will increase to a larger degree because their oxygen level will drop accordingly. It is, however, worth taking both into consideration in order to distinguish the impacts on patients’ CO_2_ and O_2_ levels, especially for patients with ventilation-perfusion inequality or right to left shunt, as their CO_2_ levels are usually normal but with low O_2_ levels.

Ventilation support will reduce patients’ work of breathing and result in less CO_2_ production because it helps drive air into the lung and lower the breathing rate if in assisted mode. Dead space ventilation will be reduced too as it is proportional to both breathing rate and dead space volume, which generally declines under moderate pressure support due to improved lung compliance and vascular resistance. Alveolar ventilation, on the other hand, will increase due to increased minute ventilation, decreased dead space ventilation, and alveolar recruitment. Therefore, the ratio of dead space ventilation to alveolar ventilation will be smaller. The state index will thus be lower and so will the state ventilation, as indicated in Eq. 7, and illustrated by both figures.

Both state ventilation and index will gradually decrease to a relatively stable value, suggesting a steady condition has been reached. Conversely, if they do not decrease as expected, the ventilation support may be ineffective in unloading the patient’s respiratory effort to reduce CO_2_ production, and/or lowering the 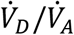 ratio for better gas exchange. In such a scenario, the doctor or physician must investigate possible causes to determine if the patient’s condition has deteriorated, or if the ventilatory support settings are inappropriate. Interpretation of the state ventilation and index must be done with care for patients who are in control modes, as they may not be reflective of respiratory effort. In fact, it is possible for patients in deep sedation that a higher number indicates the gain of conscious breathing. Hence, it is important to check the other ventilation parameters before drawing conclusions.

To investigate the root cause of ventilation failure and to facilitate clinical decision making, it is important to have a complete picture of the respiratory characteristics of a patient. Especially for patients with acute respiratory distress syndrome, this information proves useful for adjusting lung protective ventilatory settings, evaluating responses to treatments, and predicting outcomes [16]. Though a present-day ventilator may calculate airway resistance and lung compliance from observations of pressure, flow, and volume, precise measurements of alveolar ventilation and CO_2_ production cannot be obtained because the patient’s dead space is estimated. The derived formulae here provide a means to accurately compute the above ventilation information in real time during therapy. Unlike the single point calculation in the demonstration, the computation can be continuous to obtain trends to monitor disease progression. The formulae, however, are obtained with the assumption of a stable patient condition, and the computations may thus not be accurate during the unstable stage. As indicated in Eq. 13-15, even negative values are possible if a patient’s 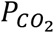 increases with an increase in ventilation or if both 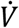 and 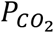 decrease simultaneously. For a patient on mechanical ventilation, the computed parameters may show oscillating behaviour before reaching a stable, accurate value, which indicates a stable condition. Therefore, the patient’s condition can be reflected by the computed trends as oscillating values may suggest an unstable condition.

A patient who reaches a very stable condition may have constant 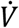 and 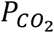, meaning that the derivatives 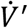 and 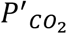 are zero (or near zero) and the applications of these equations may be impossible. In such a case, continuous computation might not be necessary since the patient’s CO_2_ production and alveolar/dead space ventilation will remain constant as well. Alternatively, the ventilator can be programmed to have its settings, such as pressure support and tidal volume, varied slowly within a small range to avoid constant 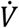 and 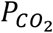, respiratory parameters can thus be easily calculated even with an assumption of linear variation.

The recent COVID-19 pandemic has led to a large increase in the demand for ventilatory support. This exacerbates the inadequacy of trained professionals in delivering complex ventilation care to their patients [17]. The challenges are not only a result of the new disease, but also arise from a generally limited understanding of ventilation parameters and outcomes.

Today, ventilators are more complicated, being equipped with many ventilation modes that each involves numerous parameters. This presents enormous challenges to trained physicians, let alone other healthcare staff. In such situations, a simple but comprehensive tool, which can evaluate the efficacy of ventilatory support, would be of significant benefit. The state ventilation, together with the other ventilation information, will provide such a tool to quantify the patient’s response and facilitate important decisions such as weaning a patient.

State ventilation and the derived formulae have other potential applications. It may be used to develop new methods to measure patient’s respiratory parameters such as dead space volume and CO_2_ production. It will enable ventilator manufacturers to develop new ventilation modes or improve the existing modes to provide better and more accurate ventilation therapies to patients. For example, the alveolar ventilation in ResMed’s iVAPS mode can be accurately computed with the presented formulae.

## CONCLUSIONS

State ventilation is a good indicator of ventilatory efficacy and a patient’s condition as it provides a measure of how much a ventilator can unload a patient’s respiratory effort and improve pulmonary gas exchange. Mathematically derived formulas can accurately and continuously compute respiratory parameters using routinely available measurements. The computations of state ventilation and respiratory parameters not only facilitate the application of AI in a ventilator device, but are also very useful for ventilation mode development, disease diagnosis, treatment selection, and outcome prediction.

## Data Availability

All data produced in the present work are contained in the manuscript

